# Evaluation of association of anti-PEG antibodies with anaphylaxis after mRNA COVID-19 vaccination

**DOI:** 10.1101/2023.04.11.23288372

**Authors:** Zhao-Hua Zhou, Margaret M. Cortese, Jia-Long Fang, Robert Wood, Donna S. Hummell, Kimberly A. Risma, Allison E. Norton, Mark KuKuruga, Susan Kirshner, Ronald L. Rabin, Cyrus Agarabi, Mary A. Staat, Natasha Halasa, Russell Ware, Anna Stahl, Maureen McMahon, Peter Browning, Panagiotis Maniatis, Shanna Bolcen, Kathryn M. Edwards, John R. Su, Sai Dharmarajan, Richard Forshee, Karen R. Broder, Steven Anderson, Steven Kozlowski

**Author notes:** Corresponding Author: Steven Kozlowski, 10903 New Hampshire Avenue, Silver Spring, MD 20993. Contributed equally (Zhao-Hua Zhou, Margaret M. Cortese).

## Abstract

**Background:** The mechanism for anaphylaxis following mRNA COVID-19 vaccination has been widely debated; understanding this serious adverse event is important for future vaccines of similar design. A mechanism proposed is type I hypersensitivity (i.e., IgE-mediated mast cell degranulation) to excipient polyethylene glycol (PEG). Using an assay that, uniquely, had been previously assessed in patients with anaphylaxis to PEG, our objective was to compare anti-PEG IgE in serum from mRNA COVID-19 vaccine anaphylaxis case-patients and persons vaccinated without allergic reactions. Secondarily, we compared anti-PEG IgG and IgM to assess alternative mechanisms.

**Methods:** Selected anaphylaxis case-patients reported to U.S. Vaccine Adverse Event Reporting System December 14, 2020 – March 25, 2021 were invited to provide a serum sample. mRNA COVID-19 vaccine study participants with residual serum and no allergic reaction post-vaccination (“controls”) were frequency matched to cases 3:1 on vaccine and dose number, sex and 10-year age category. Anti-PEG IgE was measured using a dual cytometric bead assay. Anti-PEG IgG and IgM were measured using two different assays. Laboratorians were blinded to case/control status.

**Results:** All 20 case-patients were women; 17 had anaphylaxis after dose 1, 3 after dose 2. Thirteen (65%) were hospitalized and 7 (35%) were intubated. Time from vaccination to serum collection was longer for case-patients vs controls (post-dose 1: median 105 vs 21 days). Among Moderna recipients, anti-PEG IgE was detected in 1 of 10 (10%) case-patients vs 8 of 30 (27%) controls (p=0.40); among Pfizer-BioNTech recipients, it was detected in 0 of 10 case-patients (0%) vs 1 of 30 (3%) controls (p>0.99). Anti-PEG IgE quantitative signals followed this same pattern. Neither anti-PEG IgG nor IgM was associated with case status with both assay formats.

**Conclusion:** Our results support that anti-PEG IgE is not a predominant mechanism for anaphylaxis post-mRNA COVID-19 vaccination.

## INTRODUCTION

Soon after the introduction of mRNA COVID-19 vaccines in adults in December 2020 in the United Kingdom and the United States, cases of anaphylaxis were reported, raising concerns that rates were higher than seen with other vaccines.[1] In the United States, reporting rates for anaphylaxis from the Vaccine Adverse Event Reporting System (VAERS) data through January 18, 2021 were estimated as 4.7 cases per million Pfizer-BioNTech vaccine doses administered and 2.5 cases per million Moderna vaccine doses administered.[2, 3] From the Vaccine Safety Datalink (VSD), a collaboration between CDC and 9 integrated healthcare plans with a covered population of ∼12 million people, the estimated rates of ∼5 cases per million doses for each of the vaccine products through May 2021 appeared somewhat higher than previous estimates from this system for trivalent inactivated influenza vaccine (the vaccine with the largest number of doses administered in the VSD evaluation), 1.35 cases of anaphylaxis per million doses administered.[4, 5]

Identifying the mechanism for anaphylaxis following mRNA COVID-19 vaccines is important because several doses are needed for optimal protection, likely including repeated boosters and mRNA technology is expected to be used in vaccines targeting other infectious diseases. One component shared by both mRNA COVID-19 vaccine lipid nanoparticles (LNP) is polyethylene glycol (PEG) .[6, 7] PEG has been previously identified as a potential allergen in medication reactions and thus was considered the potential cause of the post-mRNA COVID-19 vaccine anaphylaxis.[1, 8] Proposed alternatives to anti-PEG IgE-mediated type 1 hypersensitivity have included complement activation-related pseudoallergy (CARPA, which could involve IgG or IgM antibodies, with PEG as one of the proposed antigens), and direct mast cell effects through mRNA or other vaccine components.[9-13]

Detecting anti-PEG IgE among patients who had anaphylaxis after mRNA COVID-19 vaccination could identify the allergen and mechanism of these reactions. Given that individuals with a history of anaphylaxis to PEG-containing medications had been confirmed by the presence of anti-PEG IgE,[14] we anticipated that detection of anti-PEG IgE in serum of patients after anaphylaxis from mRNA COVID-19 vaccines, and not in controls, would be an indicator of its relevance. There are no widely available clinical tests for anti-PEG IgE. The accurate detection and quantitation of anti-PEG antibodies is challenging as many assay formats have high background signals and lack specificity.[15] As low levels of anti-PEG IgE may have clinical consequences, assays for anti-PEG IgE require greater sensitivity and specificity than currently possible with most assay formats. Recently a bead-based cytometry assay with internal control beads to subtract background signals was able to detect and determine titers of anti-PEG IgE in patients with PEG-associated anaphylaxis.[14] We used this assay to evaluate case-patients who had anaphylaxis after mRNA COVID-19 vaccination and control groups. Anti-PEG IgG and IgM were also evaluated using two different assay formats.

## METHODS

### Case Identification and Controls

We identified potential cases of anaphylaxis after mRNA COVID-19 vaccination, obtained and reviewed medical records, and adjudicated cases with allergists from the Clinical Immunization Safety Assessment (CISA) Project[16] to identify those most clinically consistent with anaphylaxis. Potential cases were identified through 1) searches of the passive Vaccine Adverse Event Reporting System (VAERS)[17] for reports coded as “anaphylaxis” or “anaphylactic reaction” after mRNA COVID-19 vaccination in persons aged ≥ 18 years received during December 14, 2020−March 25, 2021, and by reviews for anaphylaxis described in VAERS reports before coding, and 2) inquiries from healthcare providers or health departments to CISA. VAERS reporting for such cases was encouraged. We excluded patients with an underlying illness that could result in possible anaphylaxis (e.g. recurrent idiopathic anaphylaxis). We also classified cases by 2007 Brighton Collaboration anaphylaxis case definition levels 1-3; level 1 is the highest level of diagnostic certainty.[18, 19] Based on feasibility, our goal was to enroll ≥ 10 case-patients who had received Pfizer-BioNTech (“Pfizer-BioNTech case-patients”) and ≥ 10 who had received Moderna (“Moderna case-patients”). Case-patients were contacted via telephone and sent a project information sheet; those interested in participating provided a serum sample that was shipped to CDC laboratory. Serum was collected ≥ 6 weeks after anaphylaxis to avoid the theoretical possibility that allergen-specific serum IgE may have been reduced during the first few weeks after anaphylaxis.

Controls were selected from participants in unrelated post-authorization mRNA COVID-19 vaccine studies at 2 CISA medical centers who had serum collected under their original study protocol, provided consent for secondary use of residual serum, and for whom the specific COVID-19 vaccine dose (dose 1 vs 2) had been tolerated without allergic reaction. Serum from Pfizer-BioNTech recipients were obtained from a Vanderbilt University study [“Pfizer-BioNTech controls”], and serum from Moderna recipients [“Moderna controls”], from a Cincinnati Children’s Hospital Medical Center study. Controls were frequency matched to case-patients 3:1, by vaccine manufacturer, post-vaccine dose number (dose 1 or 2), self-identified sex and age category (10-year intervals). A control could be selected only once (i.e., to provide post-dose 1 or post-dose 2 serum). Serum provided from Pfizer-BioNTech controls had been collected ∼21 days after dose 1 (and before dose 2, if provided) and ∼21 days after dose 2; from Moderna controls, serum had been collected ∼28 days after dose 1 (and before dose 2, if provided) and ∼35 days after dose 2.

### Sample Handling and Ethics

All serum samples were first shipped to a CDC laboratory. Testing was performed in two independent laboratories. For shipment to each of the testing laboratories, separate aliquots were made and labelled with unique specimen IDs so that laboratories were blinded to case/control status and vaccine manufacturer.

This activity was reviewed by CDC and FDA in accordance with applicable regulations and institutional policies and was deemed not to be research, per 45 C.F.R.§ 46.102(l)(2), and it was determined not to be a clinical investigation as defined in 21 CFR part 56. IRB approval and formal informed consent procedures were not required.

### Anti-PEG IgE Assay

Samples were evaluated for anti-PEG IgE in the laboratory of Office of Biotechnology Products (OBP), Center for Drug Evaluation and Research (CDER), FDA using a Dual Cytometric Bead Assay (DCBA).[14] Briefly, the samples were diluted 1:5 and incubated with target beads coated with PEGylated antigen and controls beads without PEG. After washing the beads, a labeled antibody to human IgE was added. After incubation and washing, the beads were analyzed by flow cytometry. The difference in median fluorescence intensity (MFI) between the target and control beads was used to determine anti-PEG IgE antibody positivity. The assay in the current study was qualified to be specific and sensitive enough to detect ≤200 pg/mL anti-PEG IgE using a commercial anti-PEG IgE as a standard.

A cut-point for assay positivity was determined using a separate set of pre-existing sera (negative panel). These sera were commercially sourced deidentified samples used in a prior study[14] and tested negative. The negative panel and anti-PEG IgE standard run were repeated for assay consistency with three preparations of target and control beads used in the current project. For the primary objective, the cut-point was defined to be the average signal of the negative panel of pre-existing sera plus three standard deviations (SD). A sensitivity analysis was performed with a 2-SD cut-point. Examples of anti-PEG IgE assay quantitative signals are included in Supplementary Materials from the negative panel (Figure S1A) and our participant samples (Figure S1B).

A positive IgE result for a sample was defined as a signal (MFI) that is greater than the pre-defined cut-point value in two determinations and that can be inhibited by greater than 30% with the addition of free pegfilgrastim at 5 mcg/ml (demonstrating PEG specificity). Although there were positive samples based on the criteria, all of the signals were at the low end of the assay dynamic range, well below 1 ng/mL and lower than a prior sample from a patient with PEG allergy (Supplementary Figure S2). Additional details are in Supplementary Materials.

### Anti-PEG IgG and IgM assays

Anti-PEG IgG and IgM were each evaluated by flow cytometry using two different platforms in two different laboratories. The laboratories were blinded to each other’s results as well as to sample case vs. control status to minimize bias. The two assay platforms were the Dual Cytometric Bead Assay (DCBA) and a PEGylated Polystyrene Beads Assay (PPBA) that used commercially available PEGylated beads (TentaGel™ M OH beads). The DCBA was performed in OBP, CDER. The PPBA was performed in the laboratory of National Center for Toxicological Research (NCTR).

For the anti-PEG IgG and IgM DCBA, similar to the anti-PEG IgE assay above, labeled antibodies to human IgG and IgM were added after the sample incubation and washing. The titer was defined as the highest dilution of sera that led to a 100% increase in signal over the control beads. A positive result was defined as a titer of ≥ 1:20. Examples of sample titrations for the IgG dual bead method are in Supplementary Figure S2.

For the PPBA,[20] the beads were incubated with human samples diluted in 5% BSA, washed and stained for bound IgG or IgM with a specific fluorescence conjugated anti-human IgG or IgM secondary antibody, washed and analyzed by flow cytometry. The MFI was used to determine the presence or absence of anti-PEG antibodies. The cut-point for sample positivity is equal to the MFI of the negative controls (5% BSA) of each experiment plus 3 SD. The samples were also diluted, and the titer was defined as the highest dilution of sera that exceeded the cut-point for positivity.

### Statistical analysis

Fisher’s exact test was used to compare the proportion of case-patients vs controls positive for the specific anti-PEG antibody isotype (IgE, IgG, IgM), for both vaccine manufacturers combined, and for each vaccine separately. A p-value<.05 was considered statistically significant. The IgG and IgM results were assessed separately for each testing platform. No adjustment was made for multiple comparisons.

We compared the differences between means of anti-PEG IgE signal intensity of the case-patient, control and negative panel groups using the Tukey Kramer test. We determined the correlation between anti-PEG IgG results from the two assays using all case-patients and controls combined using linear regression for titers and percent concordance for positivity (Supplementary Figure S3 and Table S1). The correlation between anti-PEG IgE (positive vs negative) and anti-PEG IgG titer was assessed using Wilcoxson rank sum test. The correlation between anti-PEG IgM (positive vs negative) and anti-PEG IgG titer was similarly assessed. Analyses were performed with JMP 16 (SAS Institute, Inc. Cary, NC) and Stata 15 (College Station, TX).

## RESULTS

### Case enrollment and timing of serum sample collection

Consistent with the initial reports of allergic reactions occurring primarily in female healthcare workers, all case-patients selected to be approached for enrollment were female. Of the 25 case-patients that were reached by telephone and were provided study information, 20 participated, 1 wished to participate but was ineligible because of an underlying condition, 2 declined and for 2 follow-up calls were not returned.

The median age of the Pfizer-BioNTech and Moderna case-patients was 40.5 years and 46.5 years, respectively (Table 1). Seventeen case-patients had anaphylaxis after dose 1 and 3 after dose 2. All had onset of symptoms ≤15 minutes post-vaccination. In total, 13 of 20 (65%) were admitted to the hospital and 7 of 20 (35%) were intubated **(Table 1)**.

**Table 1.** Selected features of anaphylaxis case-patients*

| **Characteristic** | **Total**<br>**case-patients, n (%)**<br>**N=20** | **Pfizer-BioNTech**<br>**recipients, n (%)**<br>**N=10** | **Moderna**<br>**recipients, n (%)**<br>**N=10** |
| --- | --- | --- | --- |
| Female sex (self-identified) | 20 (100) | 10 (100) | 10 (100) |
| Age, median, yrs (IQR)<br>range | 43.6 (36, 52)<br>28–57 | 40.5 (32, 52)<br>39–57 | 46.5 (39, 52)<br>28–54 |
| Post dose 1 | 17 (85) | 9 (90) | 8 (80) |
| Post dose 2 | 3 (15) | 1 (10) | 2 (20) |
| Onset ≤15 minutes post-vaccination | 20 (100) | 10 (100) | 10 (100) |
| Managed in ED only | 7 (35) | 5 (50) | 2 (20) |
| Admitted to hospital or<br>observed ≥1 night | 13 (65) | 5 (50) | 8 (80) |
| Intubated | 7 (35) | 2 (20) | 5 (50) |
| Treatment included: |  |  |  |
| Epinephrine (IM or IV) | 20 (100) | 10 (100) | 10 (100) |
| Epinephrine infusion | 7 (35) | 3 (30) | 4 (40) |
| Systemic corticosteroids | 20 (100) | 10 (100) | 10 (100) |
| Brighton level |  |  |  |
| Level 1 | 14 (70) | 7 (70) | 7 (70) |
| Level 2 | 6 (30) | 3 (30) | 3 (30) |

*ED, emergency department*
\*Only 2 patients had tryptase measured in serum collected <24 hours after onset of reaction (one 2.75 hours after onset; one 7 hours after onset; both <5.5 ng/ml)

Among Pfizer-BioNTech recipients in the post-dose 1 comparison, median number of days from dose to sample collection was 105 for case-patients and 21 for controls (Supplementary Materials Table S2). Among Moderna post-dose 1 recipients, median days to sample collection was 107 vs 26 for case-patients vs controls, respectively. While receipt of dose 2 mRNA COVID-19 vaccine was not considered during selection of post-dose 1 controls, all controls who had contributed post-dose 1 serum had shortly thereafter received dose 2 without allergic reaction (one had mild asthma exacerbation within 4 weeks post-dose 2–exact onset unknown−which she considered unlikely related to vaccination).

### Anti-PEG IgE Results

Using the 3-SD cut-point for positivity, with recipients of either vaccine combined, 1 of 20 (5%) case-patients and 9 of 60 (15%) controls were positive for anti-PEG IgE (p=0.44) **(Table 2**). Among Moderna recipients, the proportion positive tended to be higher among controls vs case-patients: 8 of 20 (27%) controls vs 1 of 10 (10%) case-patients were positive (p = .40). Among Pfizer-BioNTech recipients, a single control (1 of 30, 3%) and no cases (0 of 10; 0%) were anti-PEG IgE positive (p>.99) **(Table 2)**. Results were similar when stratified by first or second vaccine doses (Supplementary Table S3).

**Table 2.** Comparison of Anti-PEG IgE Positivity in Case-patients and Controls^a^

| Category | 3 SD Threshold for Positivity |  |  | 2 SD Threshold for Positivity |  |  |
| --- | --- | --- | --- | --- | --- | --- |
|  | Case-patients<br>n (%) | Controls<br>n (%) | P-value | Case-patients<br>n (%) | Controls<br>n (%) | P-value |
| **Recipients of either vaccine** |  |  | 0.44 |  |  | 0.13 |
| Anti-PEG IgE positive | 1 (5) | 9 (15) |  | 2 (10) | 17 (28) |  |
| Anti-PEG IgE negative | 19 (95) | 51 (85) |  | 18 (90) | 43 (72) |  |
| Total | 20 | 60 |  | 20 | 60 |  |
| **Moderna vaccine recipients** |  |  | 0.40 |  |  | 0.06 |
| Anti-PEG IgE positive | 1 (10) | 8 (27)<sup>b</sup> |  | 1 (10) | 14 (47)<sup>c</sup> |  |
| Anti-PEG IgE negative | 9 (90) | 22 (73)<sup>b</sup> |  | 9 (90) | 16 (53)<sup>c</sup> |  |
| Total | 10 | 30 |  | 10 | 30 |  |
| **Pfizer-BioNTech vaccine recipients** |  |  | >.99 |  |  | >.99 |
| Anti-PEG IgE positive | 0 (0) | 1 (3)<sup>d</sup> |  | 1 (10) | 3 (10)<sup>e</sup> |  |
| Anti-PEG IgE negative | 10 (100) | 29 (97)<sup>d</sup> |  | 9 (90) | 27 (90)<sup>e</sup> |  |
| Total | 10 | 30 |  | 10 | 30 |  |

<sup>a</sup>Positivity is based on a signal greater than 3 SD above the negative panel average in two determinations and 30% inhibition by PEG-filgrastim. A sensitivity analysis with the same requirements at 2 SD above the negative panel was also performed. Fisher's exact p-values are shown above each table. Post dose 1 and post dose 2 samples are combined in this analysis.
Post-hoc unmatched analyses comparing proportion of controls anti-PEG IgE positive among Moderna vs Pfizer-BioNTech recipients: b vs d, p=.03; c vs e, p=.003

Results were in the same direction when the 2-SD cut-point was used in the sensitivity analysis, with additional samples IgE positive among controls and case-patients. For recipients of either vaccine combined, 17 of 60 (28%) controls and 2 of 20 (10%) case-patients were anti-PEG IgE positive (p=.13) **(Table 2**). As with the 3-SD cut-off, the proportion positive tended to be higher among Moderna controls vs Moderna case-patients (14 of 30 [47%] vs 1 of 10 [10%], p=.06). For Pfizer-BioNTech recipients, positivity using a 2-SD cutoff was 10% for both controls (3 of 30) and case-patients (1 of 10) (p>.99).

With a pattern similar to that of the proportion anti-PEG IgE positive using the cut-point criteria, the anti-PEG IgE quantitative signals (comparing MFI) were statistically significantly higher for controls vs case-patients among Moderna recipients, and for these controls vs the negative panel samples (Supplemental Figure S5). Among Pfizer-BioNTech recipients, there were no statistically significant differences in the quantitative anti-PEG IgE signals for case-patients vs controls, nor each group vs the negative panel samples.

In post-hoc unmatched comparison of controls by vaccine manufacturer, the proportion of Moderna controls anti-PEG IgE positive was statistically significantly higher compared with that of Pfizer-BioNTech controls (using 3-SD cut-point, 27% vs 3%, p=.03, using 2-SD cut-point 47% vs 10%, p=.003, respectively) **(Table 2)**

### Anti-PEG IgG and IgM Results

The proportion of samples anti-PEG IgG positive was higher in control vs case-patients for recipients of either vaccine (DCBA: 52% vs. 20%, p = 0.014; PPBA: 55% vs. 45%, p = 0.45) and for recipients of Moderna vaccine (DCBA: 67% vs. 20%, p = 0.003; PPBA: 67% vs. 40%, p = 0.16), although the differences were statistically significant only with DCBA **(Table 3)**. Among Pfizer-BioNTech recipients, with each assay, the proportion of controls vs case-patients anti-PEG IgG positive was similar (∼30-50%) **(Table 3)**. The general pattern is similar to that observed with the anti-PEG IgE evaluation.

**Table 3.** Comparison of Anti-PEG IgG and IgM Positivity in Case-patients and Controls^a^

| Antibody and category | DCBA Assay |  |  | PPBA Assay |  |  |
| --- | --- | --- | --- | --- | --- | --- |
|  | Case-patients<br>n (%) | Controls<br>n (%) | P-value | Case-patients<br>n (%) | Controls<br>n (%) | P-value |
| **Anti-PEG IgG** |  |  |  |  |  |  |
| **Recipients of either vaccine** |  |  | 0.01* |  |  | 0.45 |
| Anti-PEG IgG positive | 4 (20) | 31 (52) |  | 9 (45) | 33 (55) |  |
| Anti-PEG IgG negative | 16 (80) | 29 (48) |  | 11 (55) | 27 (45) |  |
| Total | 20 | 60 |  | 20 | 60 |  |
| **Moderna vaccine recipients** |  |  | 0.003* |  |  | 0.16 |
| Anti-PEG IgG positive | 1 (10) | 20 (67)<sup>b</sup> |  | 4 (40) | 20 (67)<sup>c</sup> |  |
| Anti-PEG IgG negative | 9 (90) | 10 (33)<sup>b</sup> |  | 6 (60) | 10 (33)<sup>c</sup> |  |
| Total | 10 | 30 |  | 10 | 30 |  |
| **Pfizer-BioNTech vaccine recipients** |  |  | >.99 |  |  | 0.73 |
| Anti-PEG IgG positive | 3 (30) | 11 (37)<sup>d</sup> |  | 5 (50) | 13 (43)<sup>e</sup> |  |
| Anti-PEG IgG negative | 7 (70) | 19 (63)<sup>d</sup> |  | 5 (50) | 17 (57)<sup>e</sup> |  |
| Total | 10 | 30 |  | 10 | 30 |  |
| **Anti-PEG IgM** |  |  |  |  |  |  |
| **Recipients of either vaccine** |  |  | 0.44 |  |  | 0.62 |
| Anti-PEG IgM positive | 1 (5) | 8 (13) |  | 10 (50) | 26 (43) |  |
| Anti-PEG IgM negative | 19 (95) | 52 (87) |  | 10 (50) | 34 (57) |  |
| Total | 20 | 60 |  | 20 | 60 |  |
| **Moderna vaccine recipients** |  |  | 0.17 |  |  | 0.16 |
| Anti-PEG IgM positive | 0 (0) | 8 (27)<sup>f</sup> |  | 4 (40) | 20 (67)<sup>g</sup> |  |
| Anti-PEG IgM negative | 10 (100) | 22 (73)<sup>f</sup> |  | 6 (60) | 10 (33)<sup>g</sup> |  |
| Total | 10 | 30 |  | 10 | 30 |  |
| **Pfizer-BioNTech vaccine recipients** |  |  | 0.25 |  |  | 0.04* |
| Anti-PEG IgM positive | 1 (10) | 0 (0)<sup>h</sup> |  | 6 (60) | 6 (20)<sup>j</sup> |  |
| Anti-PEG IgM negative | 9 (90) | 30 (100)<sup>h</sup> |  | 4 (40) | 24 (80)<sup>j</sup> |  |
| Total | 10 | 30 |  | 10 | 30 |  |

<sup>a</sup>Positivity is based on a titer of $\geq 1:20$ for dual bead cytometric assay (DCBA) and greater than 3SD threshold for the PEG polystyrene bead assay (PPBA). The Fisher's exact test p-values are shown above each table. Significant results of $p < 0.05$ are followed by an asterisk.
Post-hoc unmatched analyses comparing proportion of controls anti-PEG IgG positive (or anti-PEG IgM positive) among Moderna vs Pfizer-BioNTech recipients: b vs d, $p=.04$ ; c vs e, $p=.12$ ; f vs h, $p=.005$ , g vs j, $p=.001$

For anti-PEG IgM using DCBA, a relatively low proportion of participants were positive with no statistically significant differences in case-patients vs controls among Moderna recipients nor among Pfizer-BioNTech recipients **(Table 3)**. There tended to be a higher proportion of participants who were anti-PEG IgM positive by PPBA compared with DCBA. With PPBA, among Pfizer-BioNTech recipients, anti-PEG IgM positivity was higher in case-patients vs controls (6 of 10 (60%) vs 6 of 30 (20%), p=.04) **(Table 3)**.

With both assays, the anti-PEG IgG titers correlated with the anti-PEG IgE result (Supplementary Figure S6), with IgE-positive participants having higher IgG titers (for each assay, p<.001). Within each assay type, the anti-PEG IgG titers also correlated with the anti-PEG IgM result, with IgM-positive participants having higher IgG titers (for each assay type, p<.001).

In post-hoc comparison of groups by vaccine manufacturer, with DCBA, the proportion of Moderna controls anti-PEG IgG-positive was statistically significantly higher compared with Pfizer-BioNTech controls **(Table 3**). With both assay types, anti-PEG IgM positivity was statistically significantly higher in Moderna controls vs Pfizer-BioNTech controls. Actual anti-PEG IgG titers were higher among Moderna controls vs case-patients, but this was not observed for Pfizer-BioNTech recipients (Supplementary Figure S7). Anti-PEG IgG and IgM titers in Moderna controls were higher than those in Pfizer-BioNTech controls.

## DISCUSSION

IgE-mediated hypersensitivity is a potential mechanism for the observed mRNA COVID-19 vaccine anaphylaxis and PEG has been a suspected allergen. In our project with 20 patients with clinical anaphylaxis post-mRNA COVID-19 vaccination and 60 matched controls who tolerated vaccination without allergic reactions, only 1 case-patient had detectable anti-PEG IgE antibodies using a sensitive assay and there was no positive correlation between anaphylaxis case status and anti-PEG IgE antibody positivity. This was true when Moderna and Pfizer-BioNTech COVID-19 vaccine recipients were analyzed together, or when evaluating each vaccine individually.

Our results support that pre-existing anti-PEG IgE is not the mechanism for many post-mRNA COVID-19 vaccine anaphylaxis cases and is consistent with other studies and clinical observations. Additionally, the presence of anti-PEG IgE in some of our controls post-dose 1 (all of whom subsequently tolerated dose 2) suggests that the levels detected in our project were not clinically relevant. Warren et al[21] evaluated anti-PEG IgE with an ELISA assay in serum of 12 patients who met Brighton anaphylaxis criteria 1 or 2 (8 after Pfizer-BioNTech, 4 after Moderna) and none tested positive. Additional patients with immediate non-anaphylactic allergic reactions and the 3 controls assessed (2 post Pfizer-BioNTech, 1 post Moderna) were also anti-PEG IgE negative. Anti-PEG IgE was not detected in a patient described as having had a severe allergic reaction requiring hospitalization following mRNA COVID-19 vaccination.[22] However, Mouri et al[23] detected anti-PEG IgE using an ELISA assay in a patient with Brighton level 3 anaphylaxis after Pfizer-BioNTech vaccination, as well as in other patients with immediate non-anaphylactic and delayed reactions following mRNA COVID-19 vaccines. In their study, anti-PEG IgE levels in the immediate reaction group were higher than in the control group (all Pfizer-BioNTech recipients); some controls were anti-PEG IgE positive. Differences in anti-PEG assays are likely a key reason for disparity among some reports; populations and classification of reactions may also contribute. Importantly, there are several reports describing patients with anaphylaxis or suspected anaphylaxis following first dose of mRNA COVID-19 vaccines who subsequently tolerated a second dose without a serious allergic reaction.[24, 25] These patients had been carefully evaluated and monitored; many were pre-medicated for the second dose and received it with graded administration. A larger number of patients have been reported who had immediate, non-anaphylactic suspected allergic reactions following a first mRNA COVID-19 vaccine and subsequently received a second dose without a serious reaction.[24, 26] These reports of dose 2 tolerance support that at least some anaphylaxis cases are due to mechanisms other than typical IgE-mediated type 1 hypersensitivity, regardless of the exact allergen. Four anti-PEG IgE-positive patients in Mouri et al had also received a second dose of mRNA COVID-19 vaccine, and all tolerated without allergic reaction.

We also did not find a positive correlation between cases and anti-PEG IgG positivity or titers; however, anti-PEG IgM positivity was higher in case-patients than controls for Pfizer-BioNTech recipients only with the PPBA assay (60% vs 20%, p=.04 without adjusting for multiple comparisons). In contrast to our anti-PEG IgG findings, Warren et al detected anti-PEG IgG in 10 of 11 anaphylaxis case-patients but in none of the 3 controls. The difference in assays may be key. Additionally, Warren et al case-patient samples were collected sooner after anaphylaxis (median 36.5 days, range 0-78) than ours (median 105 days). Their lower proportion of systemic corticosteroid receipt (4 of 12, 33% vs our 20 of 20, 100%) may have also possibly contributed. Our control group was larger–30 per vaccine–and with samples collected a median of ∼21 days post-vaccination, we detected anti-PEG IgG in 37% – 67% depending on vaccine and assay. Lim et al reported higher levels of anti-PEG IgG or anti-PEG IgM (anti-PEG IgE not evaluated) in 2 of 3 patients with suspected anaphylaxis from Pfizer-BioNTech vaccine vs controls.[27]

We found some differences by vaccine type. Moderna controls tended to have higher frequency and signal intensity of anti-PEG IgE compared with Moderna case-patients or Pfizer-BioNTech controls. Moderna controls had a higher frequency of anti-PEG IgG (DCBA) and IgM (both assays) than Pfizer-BioNTech controls. Moderna controls also had higher titers of anti-PEG IgG and IgM than Pfizer-BioNTech controls with both assay formats. Other studies have reported boosting of anti-PEG IgG in Moderna vs Pfizer-BioNTech recipients who tolerated vaccination.[22, 28]

Strengths of our evaluation include cases selected from reports to national VAERS and CISA infrastructures and adjudicated by CISA allergists, matched controls at a 3:1 ratio, blinding of samples, and use of two independent assays when feasible. Limitations include case-patient sample size based on feasibility. As with all such retrospective case reviews, particularly if tryptase was not measured during the recommended period to aid in the assessment, it is possible that at least some of our cases were not anaphylaxis. Most of our case-patients were treated promptly with epinephrine which may have ameliorated severity. However, anaphylaxis can be very challenging to differentiate from other immediate non-allergic reactions, including vocal cord dysfunction, immunization stress-related response, and vasovagal reactions.[29-31] The shorter time to sample collection among our controls vs case-patients may explain the higher frequencies and titers for some anti-PEG “non-mechanistic” antibodies; the receipt of corticosteroids by all our cases may also have contributed. Other possible limitations include assay performance, the nature of the PEG antigen (e.g., PEGylated lipids)[32] and mast-cell associated anti-PEG antibodies not being reflected in serum levels.

Although we cannot exclude typical anti-PEG IgE-mediated type 1 hypersensitivity as a mechanism for mRNA COVID-19 vaccine anaphylaxis, these results add further doubt as to this being the predominant mechanism.

## Data Availability

Patient specific data may be confidential. Other data can be shared upon request to authors.

## Acknowledgments

We would like to acknowledge the efforts of Laura Youngblood, MPH and Tom Shimabukuro, MD, MPH (CDC, Atlanta, GA), Pollyanna Stewart, MSN, RN-BC (Alaska Department of Health, Anchorage, AK), Juventila Liko, MPH and Paul Cieslak, MD (Oregon Public Health Division, Portland, OR), Frederick Little, MD (Boston Medical Center, Boston, MA), Joshua Milner, MD, PhD (Columbia University Irving Medical Center, New York, NY), Rendie McHenry, Joan Eason, Marcia Blair, and Danya Waqfi (Vanderbilt University School of Medicine), all the hospital and public health staff who assisted with sample collection, and all the participants who were willing to take part in this project.

## Financial support

This work was supported by the Centers for Disease Control and Prevention Clinical Immunization Safety Assessment (CISA) Project contracts 200-2012-53661 to Cincinnati Children’s Hospital Medical Center, 200-2012-50430 to Vanderbilt University Medical Center and 200-2012-53664 to Johns Hopkins University.

## Disclaimer

The findings and conclusions in this report are those of the authors and do not necessarily represent the official position of the CDC or the FDA. Mention of a product or company name is for identification purposes only and does not constitute endorsement by the CDC or the FDA.

## Conflict of Interest Statements

Donna S. Hummel: eDMC monitoring clinical trial (Merck)

Mary A. Staat: funding from NIH, CDC, Pfizer and Merck and royalties from UPToDate

Kathryn M. Edwards: Grant funding from NIH and CDC; Consultant to Bionet and IBM; Member Data Safety and Monitoring Board for Sanofi, X-4 Pharma, Seqirus, Moderna, Pfizer, Merck, Roche, Novavax, Brighton Collaboration

The other authors have no conflicts of interest.

## Notes

### Author Declarations

This activity was reviewed by CDC and FDA in accordance with applicable regulations and institutional policies and was deemed not to be research, per 45 C.F.R. 46.102(l)(2), and it was determined not to be a clinical investigation as defined in 21 CFR part 56. IRB approval and formal informed consent procedures were not required.

