## Supplementary Materials for "Evaluation of association of anti-PEG antibodies with anaphylaxis after mRNA COVID-19 vaccination"

### Supplementary Materials Expanded Methods

#### *Case Enrollment*

After cases were selected, health department personnel from the case-patients' jurisdictions were contacted to determine the best method to contact the patients and request participation. Health department staff or CDC staff contacted patients via telephone to explain the project; if interested in participating, a short survey was administered to determine eligibility (i.e. patients were excluded if they had an underlying illness associated with possible anaphylaxis or if they had received a subsequent dose of mRNA COVID-19 vaccine after the anaphylaxis).

#### *Anti-PEG IgE, IgG and IgM evaluations with the Dual Cytometric Bead Assay (DCBA)* *(Referred to as Lab 1 Assay in Supplementary Materials)*

*Reagents*
Functional CBA beads E4 and E8, 7.5 µm polystyrene beads with different color-coding to differentiate bead populations, were purchased from BD Biosciences (San Jose, CA). Mouse anti-PEG IgG monoclonal antibody was purchased from Life Diagnostics, Inc. (West Chester, PA). Human Fc chimeric anti-PEG IgG and anti-PEG IgE monoclonal antibody standards were purchased from AmCell Biosciences, LLC (Mountainview, CA). Anti-human-IgE-PE monoclonal antibody was purchased from BioLegend (San Diego, CA). Anti-human IgG-PE, anti-human IgM-PE monoclonal antibodies were purchased from BD Biosciences. The specificity of these reagents was validated by flow cytometry. Pegfilgrastim (Neulasta) and pegloticase (Krystexxa) were purchased through WEP Clinical (Morrisvile, NC).

*Preparation of PEG-binding target beads and control beads and anti-PEG antibody detection by Dual Cytometric Bead Assay (DCBA)*
High affinity murine anti-PEG monoclonal antibody-conjugated functional CBA beads were prepared using sulfo-SMCC chemistry with Functional Bead Conjugation Buffer Set (BD Biosciences) according to manufacturer's instructions. Conjugation was confirmed by flow cytometry using goat anti-mouse IgG-PE and the signal to background fluorescence intensity increased by more than 10,000. CBA functional beads E4 and E8 were conjugated with the anti-PEG monoclonal antibody following the exact same procedure. E4 beads were incubated with pegloticase or other PEG-containing products in a PBS buffer containing 2mM EDTA and 1% BSA for 1 hour at room temperature with shaking, followed by washing to remove unbound pegloticase. E4 beads with a captured PEG product were defined as target beads. E8 beads conjugated with the same anti-PEG antibody but without a captured PEG product were used as control beads (Zhou ZH JACI 2021, 9:1731). The beads were stored at 4 °C and were stable for two weeks.

*Detection for anti-PEG IgG, IgM and IgE in patient sera*
Screening for anti-PEG antibodies in serum or plasma samples were carried out using 5 mL Falcon flow tubes or 96 well U-shaped plates for high-throughput runs. For every 96-well plate, ~ $10^6$ target and $10^6$ control beads were added to 2 mL serum enhancement buffer (supplied in the BD human CBA kits). Target beads and

control beads were mixed in total of 10mL buffer (4 mL serum treatment buffer, 4 mL beads capture buffer, 2 mL sample diluent). Then 100µL beads and 100 µL of diluted serum sample were added per well (96-U plate), incubated overnight at 4$^{o}$C with shaking and washed twice. Five µL/well of anti-human IgE-PE, 20 uL anti-human IgGFc-PE or 20 uL anti-human-IgM-PE were added in 100 µL buffer per well, incubate with gentle shaking for 1 hour, the wells were then washed twice and analyzed by flow cytometry using the high throughput auto sample run. Anti-PEG-IgE positive samples that had been detected positive previously by DCBA with clinical anaphylaxis symptoms to PEG were used as a positive control. Commercial positive and negative controls (AmCell Biosciences, CA) were used in some experiments. Patient plasma or serum samples from cases, controls as well as controls were evaluated for anti-PEG antibodies. Positive sera were serially diluted to determine antibody titers and specificity was confirmed by competition with free PEG (pegfilgrastim, 5 mcg/mL). Bead populations were gated by FSC-SSC. Target beads and control beads were separated by APC fluorescence intensity. PE fluorescence (antibody detection signal) was compared between target and control beads.

*Impact of anti-PEG IgG on anti-PEG IgE detection*

To test if anti-PEG IgG can mimic anti-PEG IgE signals in the DCBA assay, different concentrations (i.e., 0.97, 1.95, 3.9, 7.8, 15.6, 31.25, 62.5, 125, 250, 500 and 1000 ng/mL) of specific human anti-PEG IgG standard antibody were incubated with target (signal in blue bar) and control (signal in orange bar) bead mix and then incubated with a PE-labeled specific anti-human IgE antibody. The results in figure below showed anti-PEG IgG does not lead to an anti-PEG IgE signal (i.e., there were no meaningful difference of MFI between target and control bead populations) in the DCBA assay even with anti-PEG IgG levels of 1 mcg/mL. As a positive control (far right), 5ng/mL of anti-PEG IgE increased the target bead signal as compared to the control beads.

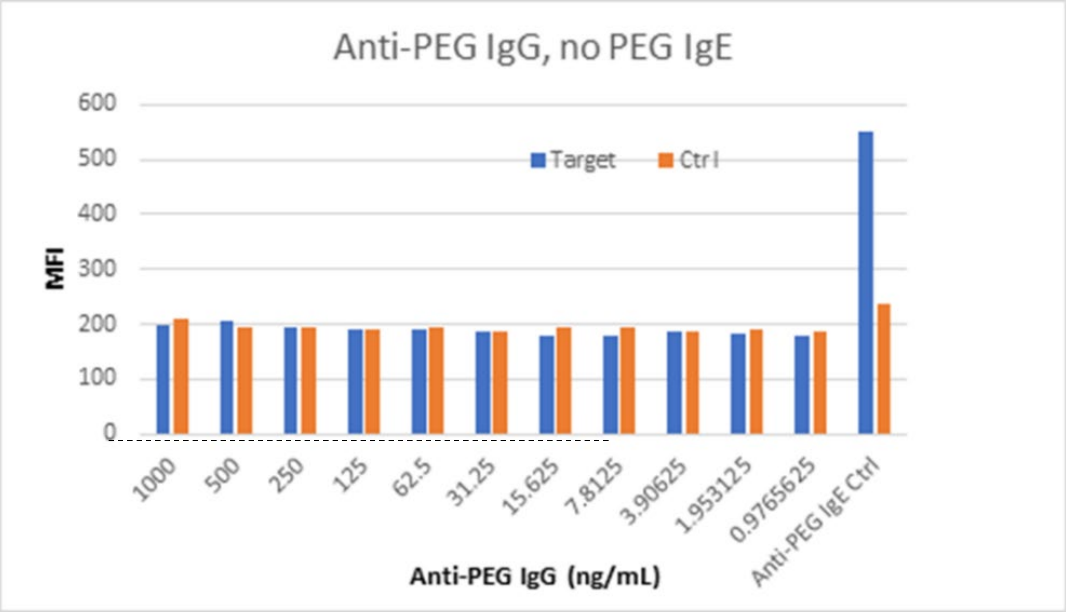

To test if anti-PEG IgG in the positive samples may interfere anti-PEG IgE signals, 2ng/mL anti-PEG IgE was mixed with different concentration of anti-PEG IgG (i.e., 1000, 500, 250, 125, 62.5, 31.25, 15.6, 7.8, 3.9, 2, 1 ng/mL) with a corresponding IgG/IgE ratio of (500, 250, 125, 62.5, 31.25, 15.6, 7.8, 3.9, 2, 1, 0.5, respectively) and evaluated for IgE signal changes by DCBA. As shown in the figure below  the anti-PEG IgE signal is only decreased with a large excess of anti-PEG IgG. BF (far right) is buffer only without anti-PEG antibody. As compared to BF, an IgG/IgE ratio of 500 still showed positive sign of anti-PEG IgE signal.

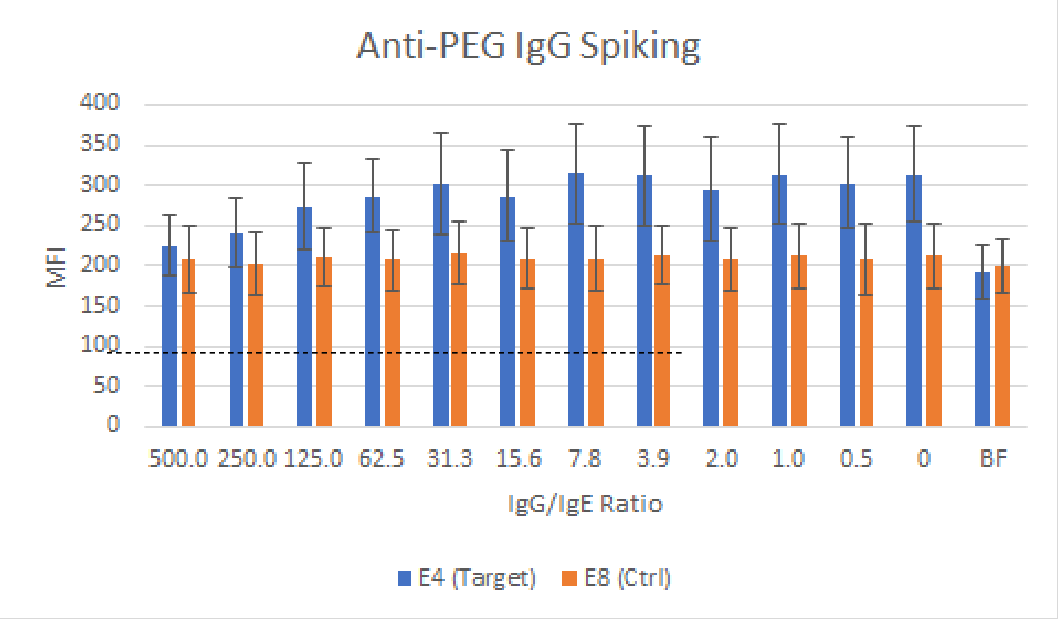

#### *Anti-PEG IgG and IgM evaluation with the PEGylated Polystyrene Beads Assay (PPBA)*
***(Referred to as Lab 2 Assay in Supplementary Materials)***

The assays were based on the publication: Fang JL, Beland FA, Tang Y, Roffler SR. Flow cytometry analysis of anti-polyethylene glycol antibodies in human plasma. Toxicol Rep. 2020 Dec 26;8:148-154. doi: 10.1016/j.toxrep.2020.12.022. PMID: 33437656; PMCID: PMC7787990.

*Reagents and antibodies:*
Human chimeric anti-PEG antibodies cHu6.3-IgG and cAGP4-IgM as standards
TentaGel™ M OH beads (particle size 10 µm)
Alexa Fluor 488 AffiniPure donkey anti-human IgM (µ-chain specific) secondary antibody
Alexa Fluor 647 AffiniPure donkey anti-human IgG (γ-chain specific) secondary antibody

*Assay:*
A 1% (w/v) stock suspension of TentaGel™ M OH beads (Lot # 1323750 12307245) was prepared in PBS without sonication.  Aliquots (25 µL) of the TentaGel™ M OH beads stock were incubated with diluted human plasma (10 µL plasma and 40 µL 5% BSA) and 100 µL of PBS by shaking at 800 rpm with an Eppendorf Thermomixer R (Eppendorf North America, Hauppauge, NY) for 1 h at room temperature. The beads were then be washed three times with PBS, and further stained for bound IgG or IgM with a specific fluorescence conjugated anti-human IgG (2.5 µL into 500 µL of PBS) or IgM secondary antibody (2.5 µL into 500 µL of PBS)

by shaking at 800 rpm for 1 h at room temperature. The stained beads were washed three times with PBS and analyzed on a Beckman Coulter CytoFlex flow cytometry (Indianapolis, IN). Data were acquired and analyzed using CytExpert 1.2 Software (Beckman Coulter). The forward-scatter (FSC) signal and the side-scatter (SSC) signal were measured in the linear mode, with a gain of 20 for FSC and 30 for SSC. Fluorescence was detected on a logarithmic scale. A total of 20,000 beads were analyzed for each sample. The median fluorescence intensity (MFI) was used to analyze the presence or absence of anti-PEG antibodies in plasma. cHu6.3-IgG (1:500) and cAGP4-IgM (1:500) were the positive anti-PEG antibody standards and 5% BSA was used as a negative control. All wash and incubation steps were performed using PBS without any surfactant.

After the complete analysis of all serum/plasma samples, a cutoff-point was established. The cutoff-point was equal to the MFI of the negative control (5% BSA) of each experiment plus 3 standard deviations, which were calculated from the MFI of the negative controls for all the experiments. The serum/plasma was considered positive for anti-PEG IgG or IgM if the MFI in serum/plasma from a serum/plasma sample was higher than the established cutoff-point.

Both cHu6.3-IgG and cAGP4-IgM standard curves were constructed by plotting the MFI versus the antibody concentration. The relative concentration of anti-PEG IgG and IgM was calculated by comparison to the cHu6.3-IgG or cAGP4-IgM standard curves, respectively.

#### Supplementary Results

Within each vaccine type, stratified analysis accounting for the frequency matching by age group yielded the same conclusions as when all age groups were combined.

#### Supplementary Figure S1

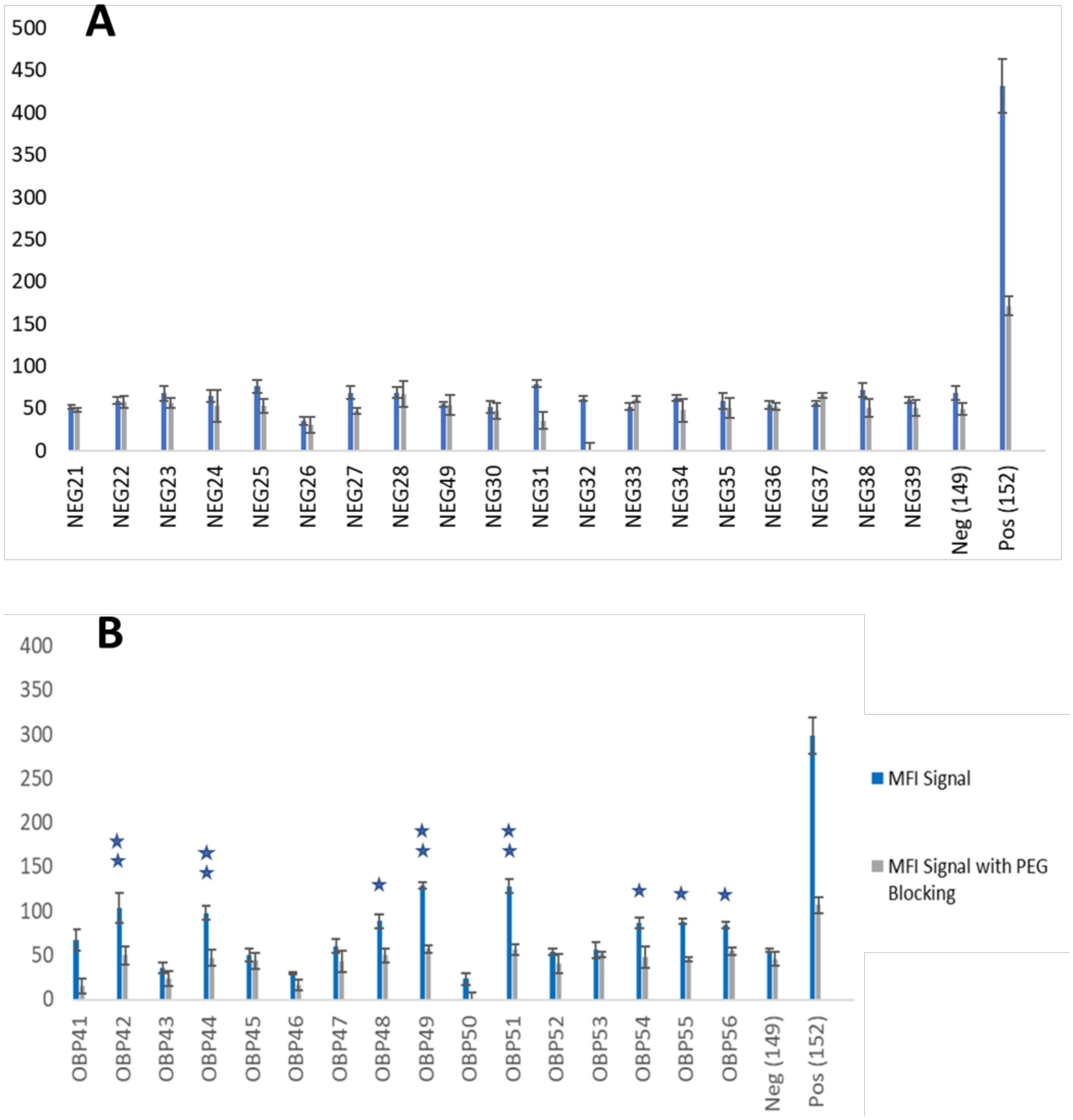

*Figure S1. Example testing of samples and controls for anti-PEG IgE positivity.*

(A) shows the MFI signal difference (MFI PEG target bead - MFI control bead) of a panel of pre-existing negative sera prior to COVID 19. The blue bars are the target and control MFI signal difference, and the grey bars are the MFI signal difference in the presence of excess PEG (pegfilgrastim) to block the PEG-specific component of the MFI signal. The MFI signal difference of the negative sera panel was used to generate the threshold for positivity for our project samples. (B) shows a representative of DCBA assay of blinded samples with OBP (Lab1) internal codes and the MFI signals were evaluated in the same way as in A. Two stars over a sample indicates a signal 3 standard deviations over the average of the negative panel. One star over a sample indicates a signal 2 standard deviations over the average of the negative panel. In the assay, code 149 was a negative control serum from a patient with a reaction after mRNA COVID-19 vaccination previously tested by DCBA and anti-PEG IgE negative and code 152 was a positive control sample from a patient with known PEG allergy (control samples code 149 and 152 were provided by Dr. Elizabeth J. Phillips, Vanderbilt University Medical Center).

#### Supplementary Figure S2

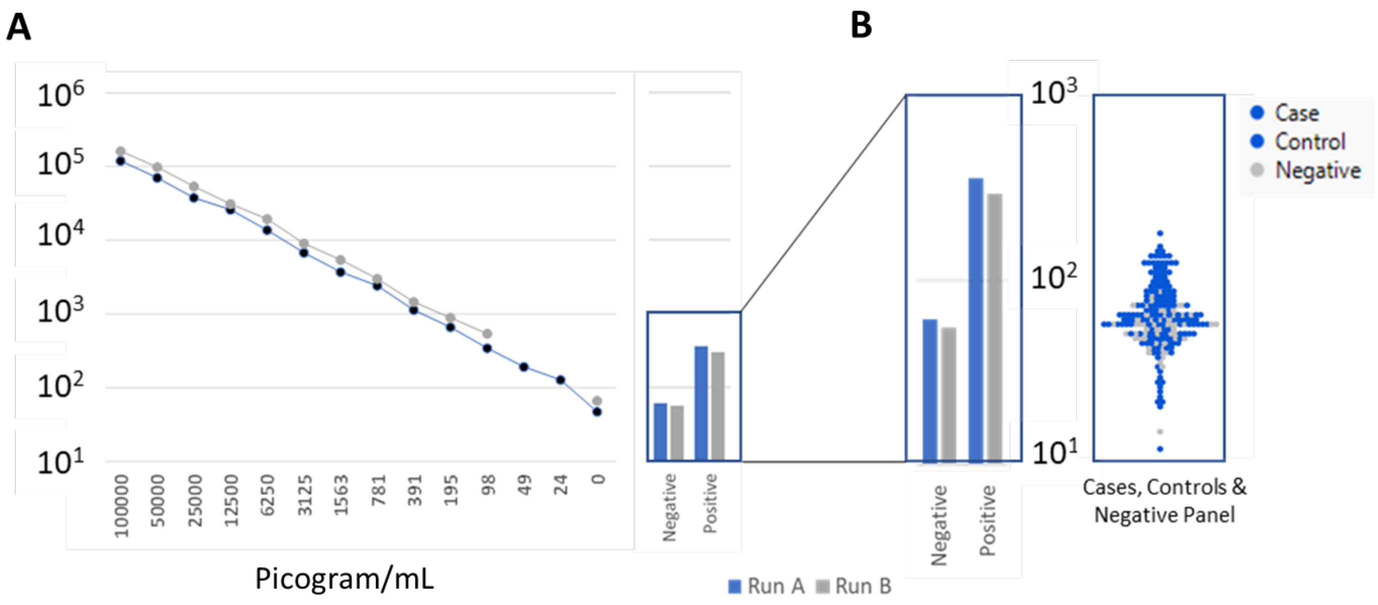

*Figure S2. Standard runs and the overall range of sample signals for the anti-PEG IgE evaluation.*

(A) Two titration runs of a monoclonal anti-PEG IgE antibody using the dual bead method are shown. The bead preparation used is the same used for the experiments in Figure 1. A positive sample from a PEG-allergic patient and a negative control are shown for each of the runs in the box. (B) The control samples in the box are enlarged and shown in comparison with all the determinations for IgE anti-PEG signals in these studies. Each determination is generally the average of 4 replicates. There were 2 determinations per test sample (vaccinated case-patients and controls) and 3 determinations per negative panel sample. Further analyses are done as per the methods.

#### Supplementary Figure S3

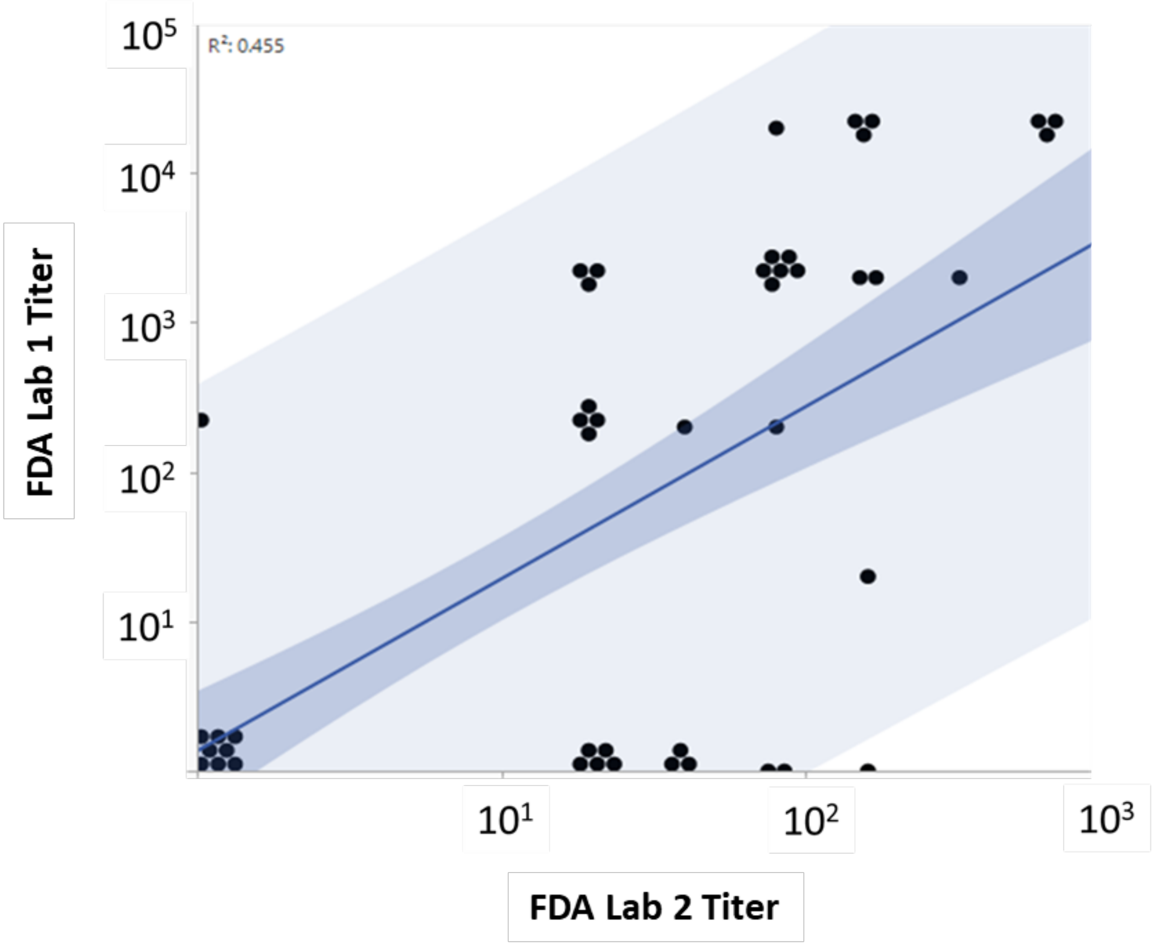

*Figure S3. Comparing Anti-PEG IgG results from Laboratory 1 and Laboratory 2*

Laboratory 1: dual cytometric bead assay (DCBA)     Laboratory 2: PEGylated polystyrene beads assay (PPBA)

The titers from FDA Laboratory 1 are graphed against the titers from FDA Laboratory 2 using all case-patients and controls combined. A linear fit with confidence and prediction intervals is shown.

#### Supplementary Figure S4

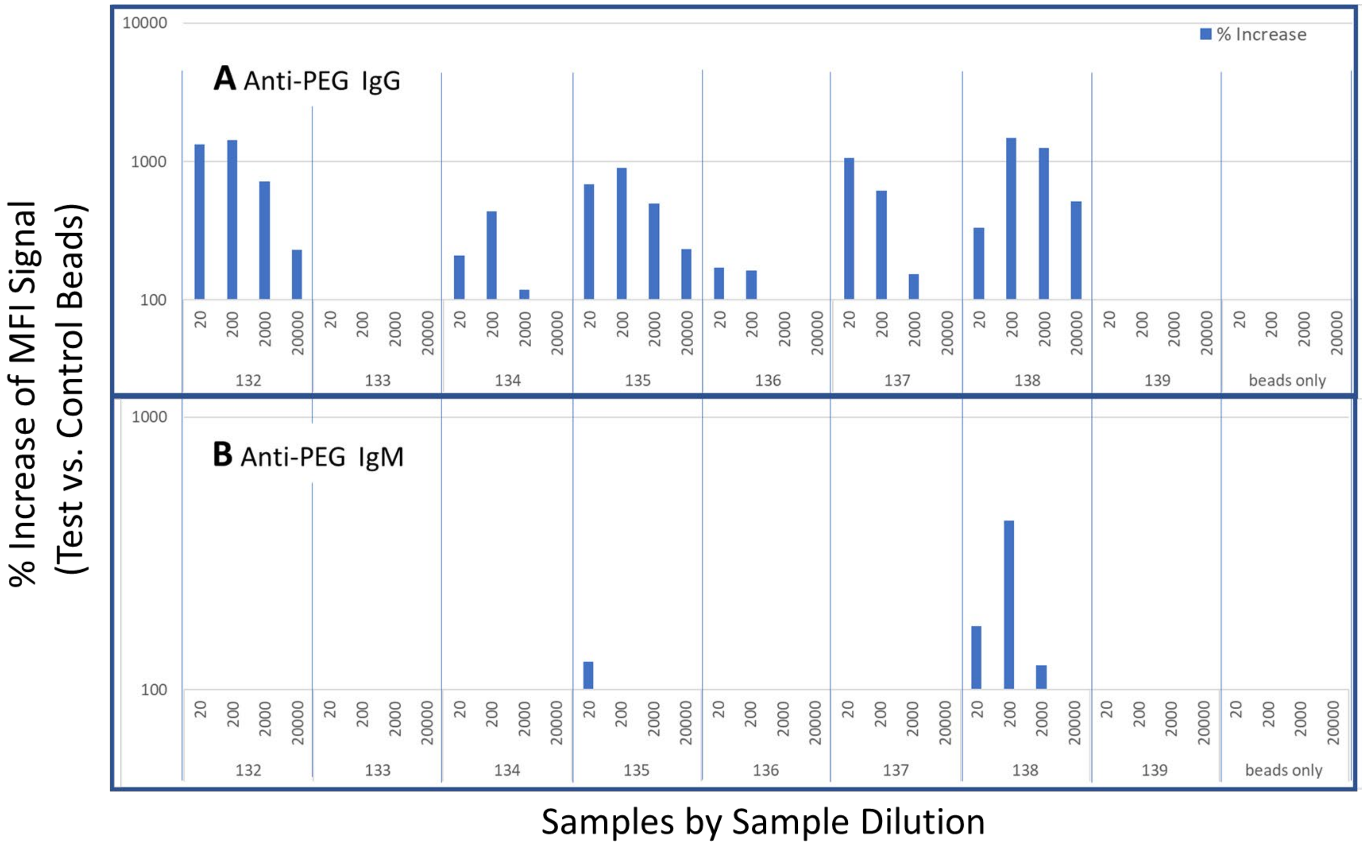

*Figure S4. Example titrations of samples to detect anti-PEG IgG (A) and anti-PEG IgM (B) using the dual bead assay.*

The Y-axis is the % increase in MFI signal of a sample with test beads (PEG) as compared to the same sample with MFI signal control beads (no PEG). The titer is the highest dilution with an MFI signal with the test bead that is 100% higher than the signal with the internal control beads.

#### Supplementary Table S1

| Table S1. Agreement between Lab 1 and Lab 2 Results[a] | | | |
|---|---|---|---|
| | Lab 1 Positive n(%) | Lab 1 Negative n(%) | Concordant Results n(%) |
| **Anti-PEG IgG** | | | 57/80 (71) |
| Lab 2 Positive | 27 (77) | 15 (33) | |
| Lab 2 Negative | 8 (23) | 30 (67) | |
| **Anti-PEG IgG with Higher Titers** | | | 64/80 (80) |
| Lab 2 Positive | 17 (81) | 12 (20) | |
| Lab 2 Negative | 4 (19) | 47 (80) | |
| **Anti-PEG IgM** | | | 53/80 (66) |
| Lab 2 Positive | 9 (100) | 27 (38) | |
| Lab 2 Negative | 0 (0) | 44 (62) | |

Laboratory 1: dual cytometric bead assay (DCBA)     Laboratory 2: PEGylated polystyrene beads assay (PPBA)

[a]The numbers of positive and negative samples using both laboratory methods are compared for anti-PEG IgG, anti-PEG IgG where positivity is defined with a higher threshold titers (1:200 or greater in Lab 1; 1:80 or greater in Lab 2), and anti-PEG IgM.

### Supplementary Table S2

| **Table S2 Timing of Serum Sample Collection** | | | | |
|---|---|---|---|---|
| | **Pfizer-BioNTech recipients** | | **Moderna recipients** | |
| | Case-patients, N | Controls, N | Case-patients, N | Controls, N |
| **Post-dose 1** | **9** | **27** | **8** | **24** |
| Days from dose to sample collection, median (IQR) | 105 (97, 128) | 21 (21, 21) | 107 (94, 132) | 26 (21,27) |
| range | 96–165 | 20–28 | 61–168 | 17–30 |
| **Post-dose 2** | **1** | **3** | **2** | **6** |
| Days from dose to sample collection | 59 | 21, 29, 31 | 77, 90 | 34 (median)<br>28-41 (range) |

#### Supplementary Table S3

| Table S3. Comparison of Anti-PEG IgE Positivity in Case-patients and Controls by Vaccine Dose[a] | | | | | | |
|---|---|---|---|---|---|---|
| | 3-SD Threshold for Positivity | | | 2-SD Threshold for Positivity | | |
| | **Cases n(%)** | **Controls n(%)** | **P-value** | **Cases n(%)** | **Controls n(%)** | **P-value** |
| **Recipients of either vaccine post-dose 1** | | | 0.67 | | | 0.32 |
| Anti-PEG IgE positive | 1 (6) | 7 (14) | | 2 (12) | 14 (27) | |
| Anti-PEG IgE negative | 16 (94) | 44 (86) | | 15 (88) | 37 (73) | |
| Totals | 17 | 51 | | 17 | 51 | |
| **Recipients of either vaccine post-dose 2** | | | >.99 | | | 0.51 |
| Anti-PEG IgE positive | 0 (0) | 2 (22) | | 0 (0) | 3 (33) | |
| Anti-PEG IgE negative | 3 (100) | 7 (78) | | 3 (100) | 6 (67) | |
| Totals | 3 | 9 | | 3 | 9 | |
| **Recipients of Moderna vaccine post-dose 1** | | | 0.64 | | | 0.10 |
| Anti-PEG IgE positive | 1 (12) | 7 (29) | | 1 (12) | 12 (50) | |
| Anti-PEG IgE negative | 7 (88) | 17 (71) | | 7 (88) | 12 (50) | |
| Totals | 8 | 24 | | 8 | 24 | |
| **Recipients of Moderna vaccine post-dose 2** | | | >.99 | | | >.99 |
| Anti-PEG IgE positive | 0 (0) | 1 (17) | | 0 (0) | 2 (33) | |
| Anti-PEG IgE negative | 2 (100) | 5 (83) | | 2 (100) | 4 (67) | |
| Totals | 2 | 6 | | 2 | 6 | |
| **Recipients of Pfizer BioNT vaccine post-dose 1** | | | >.99 | | | >.99 |
| Anti-PEG IgE positive | 0 (0) | 0 (0) | | 1 (11) | 2 (7) | |
| Anti-PEG IgE negative | 9 (100) | 27 (100) | | 8 (89) | 25 (93) | |
| Totals | 9 | 27 | | 9 | 27 | |
| **Recipients of Pfizer BioNT vaccine post-dose 2** | | | >.99 | | | >.99 |
| Anti-PEG IgE positive | 0 (0) | 1 (33) | | 0 (0) | 1 (33) | |
| Anti-PEG IgE negative | 1 (100) | 2 (67) | | 1 (100) | 2 (67) | |
| Totals | 1 | 3 | | 1 | 3 | |

[a]Positivity is based on a signal greater than 3-SD above the negative panel average in two determinations and 30% inhibition by PEG-filgrastim. A sensitivity analysis with the same requirements at -2SD above the negative panel was also performed. Fisher's exact test p-values are shown above each table.

#### Supplementary Figure S5

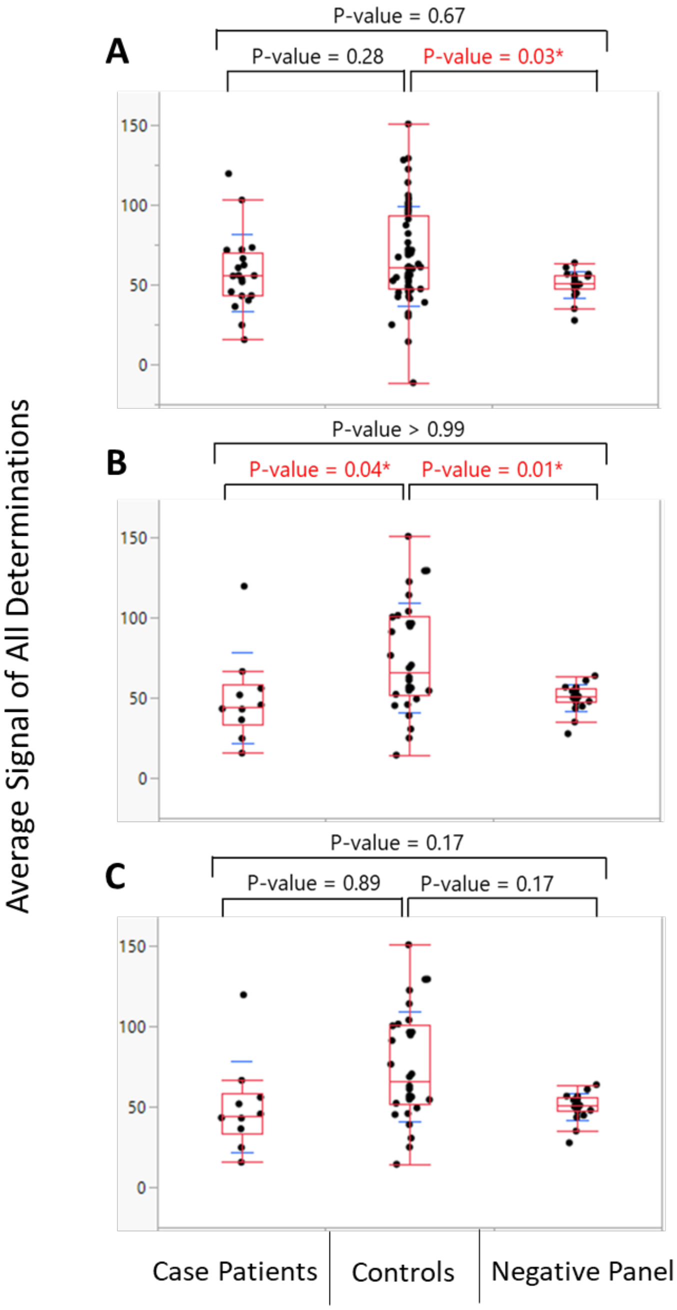

*Figure S5. Comparison of the anti-PEG IgE signal distributions between case-patients, controls and the negative panel.*

The distribution of average signal determinations is shown for all (A), Moderna (B), and Pfizer-BioNTech (C) mRNA vaccine recipients. Case-patient and control sample values are the average of two separate determinations and the negative panel samples are the average of three separate determinations. Box plots are in red and standard deviations around the mean are in blue. The points are jittered. Tukey Kramer test is used for comparing the distributions and significant differences are in red and followed by an asterisk.

#### Supplementary Figure S6

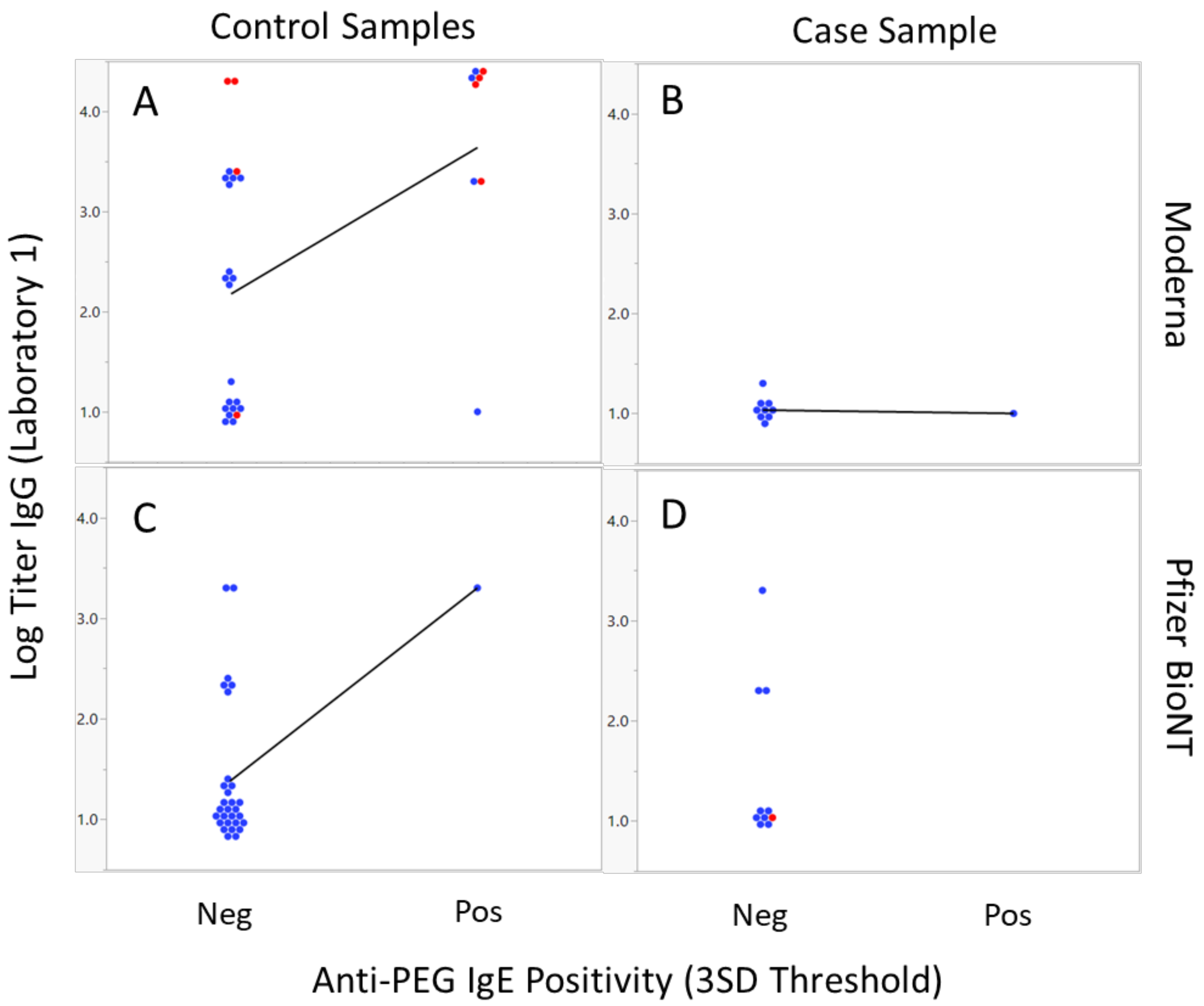

*Figure S6. Anti-PEG IgG level (and anti-PEG IgM positivity) by anti-PEG IgE positivity from Laboratory 1*

Laboratory 1: dual cytometric bead assay (DCBA)

The log titer of anti-PEG IgG from Laboratory 1 is displayed by anti-PEG IgE positivity using the 3 SD threshold. Red markers indicate the sample is also anti-PEG IgM positive; Blue markers indicate the samples was anti-PEG IgM negative. Moderna vaccine recipient control and case samples are shown in Panels A and B; Pfizer BioNTech vaccine recipient control and case samples are shown in Panels C and D. A linear fit is shown where there are both anti-PEG IgE negative and positive results.

### Supplementary Figure S7

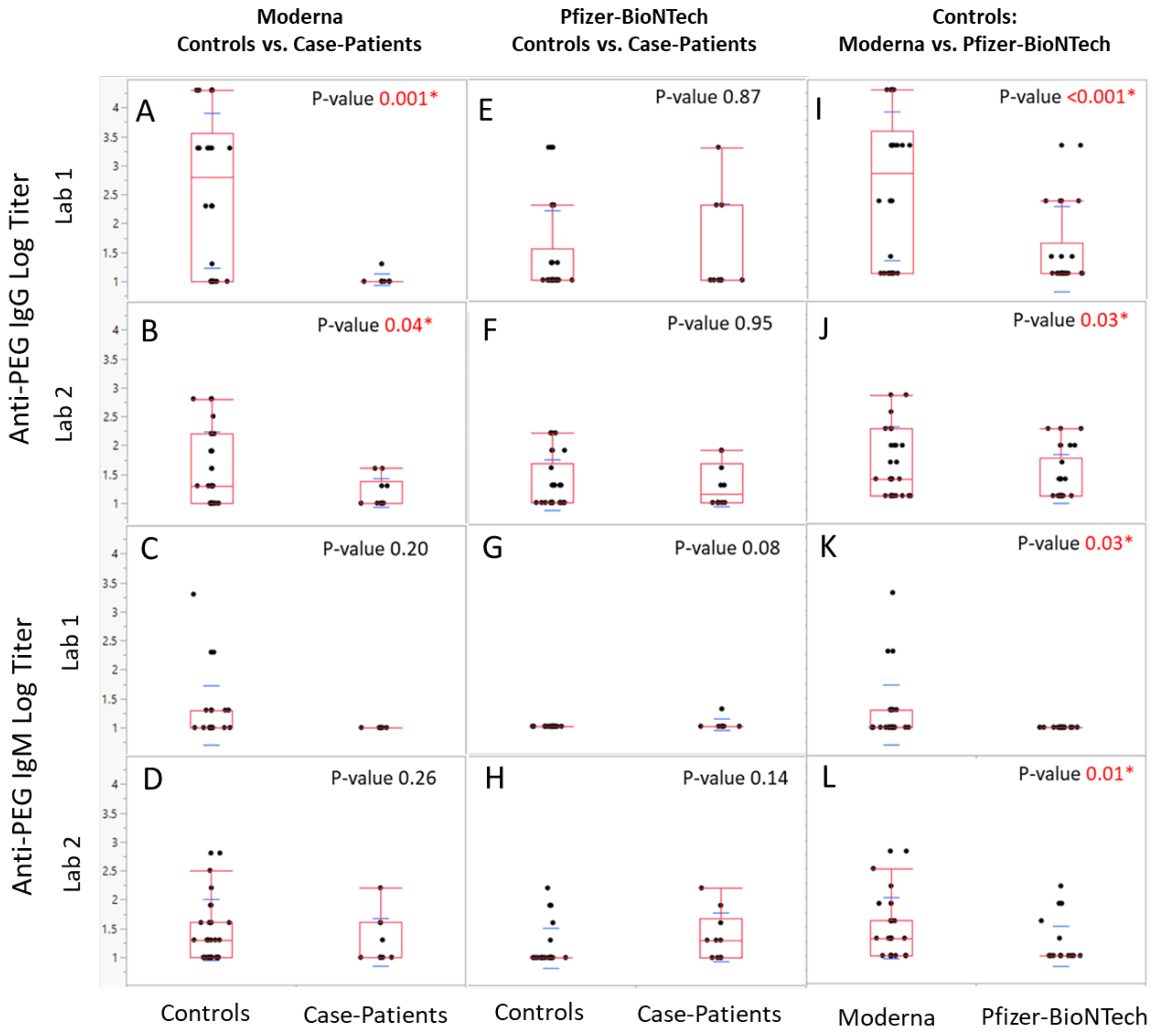

*Figure S7. Log Titers of anti-PEG IgG and anti-PEG IgM*

Laboratory 1: dual cytometric bead assay (DCBA)  Laboratory 2: PEGylated polystyrene beads assay (PPBA)

The sample titers are shown with box plots in red and standard deviations around the mean in blue. Comparisons are made using a T-test and p-values < 0.05 are shown in red with an asterisk. Titers below 20 were defined as 10 in this analysis. The displayed points are jittered. Moderna controls and case-patients are compared for anti-PEG IgG (Lab 1 assay Panel A, Lab 2 assay Panel B) and IgM (Lab 1 assay Panel C, Lab 2 assay Panel D). Pfizer-BioNTech controls and case-patients are compared for anti-PEG IgG (Lab 1 assay Panel E, Lab 2 assay Panel F) and IgM (Lab 1 assay Panel G, Lab 2 assay Panel H). Moderna and Pfizer-BioNTech controls are compared for anti-PEG IgG (Lab 1 assay Panel I, Lab 2 assay Panel J) and IgM (Lab 1 assay Panel K, Lab 2 assay Panel L)
